# Effect of Higher-Dose Fluvoxamine vs Placebo on Time to Sustained Recovery in Outpatients with Mild to Moderate COVID-19: A Randomized Clinical Trial

**DOI:** 10.1101/2023.09.12.23295424

**Authors:** The Accelerating COVID-19 Therapeutic Interventions and Vaccines (ACTIV)-6 Study Group and Investigators, Susanna Naggie

**Affiliations:** Duke Clinical Research Institute, Duke University School of Medicine, 300 West Morgan St, Suite 800, Durham, NC 27701

## Abstract

**Background:** The impact of fluvoxamine in reducing symptom duration among outpatients with mild to moderate coronavirus disease 2019 (COVID-19) remains uncertain. Our objective was to assess the effectiveness of fluvoxamine 100 mg twice daily, compared with placebo, for treating mild to moderate COVID-19.

**Methods:** The ACTIV-6 platform randomized clinical trial aims to evaluate repurposed medications for mild to moderate COVID-19. Between August 25, 2022, and January 20, 2023, 1175 participants were enrolled at 103 US sites for evaluating fluvoxamine; participants were age ≥30 years with confirmed SARS-CoV-2 infection and ≥2 acute COVID-19 symptoms for ≤7 days. Participants were randomized to receive fluvoxamine 50 mg twice daily on day 1 followed by 100 mg twice daily for 12 additional days or to placebo. The primary outcome was time to sustained recovery (defined as at least 3 consecutive days without symptoms). Secondary outcomes included time to death; time to hospitalization or death; a composite of hospitalization, urgent care visit, emergency department visit, or death; COVID clinical progression scale; and difference in mean time unwell.

**Results:** Among participants who were randomized and received study drug, the median age was 50 years (IQR 40-60), 66% were female, 45% identified as Hispanic/Latino, and 77% reported ≥2 doses of a SARS-CoV-2 vaccine. Among 589 participants who received fluvoxamine and 586 who received placebo, differences in time to sustained recovery were not observed (adjusted hazard ratio [HR], 0.99 [95% credible interval, 0.89-1.09; P(efficacy) = 0.4]). Additionally, unadjusted, median time to sustained recovery was 10 days (95% CI 10-11) in both the intervention and placebo group. No deaths were reported. Thirty-five participants reported healthcare utilization events (*a priori* defined as death, hospitalization, emergency department/urgent care visit); 14 in the fluvoxamine group compared with 21 in the placebo group (HR 0.69; 95% CrI 0.27–1.21; P(efficacy)=0.86) There were 7 serious adverse events in 6 participants (2 with fluvoxamine and 4 with placebo).

**Conclusions:** Among outpatients with mild to moderate COVID-19, treatment with fluvoxamine does not reduce duration of COVID-19 symptoms.

**Trial Registration:** ClinicalTrials.gov (NCT04885530).

## INTRODUCTION

Several clinical trials have studied approved medications as repurposed oral therapies for outpatients with mild to moderate COVID-19.^1,2^ Fluvoxamine, a selective serotonin reuptake inhibitor, has been proposed to decrease the host inflammatory response and prevent progression to severe COVID-19.^3^ A systematic review and meta-analysis suggested that fluvoxamine reduced hospitalization rates among adults with symptomatic COVID-19; however, this evidence was insufficient for national guidelines to recommend its use.^4,5^ Fluvoxamine at doses of 100 mg 2 or 3 times daily have demonstrated a reduction in emergency department visits and hospitalizations,^5,6^ although tolerability may be a limitation. A lower dose of 50 mg twice daily had improved tolerability,^6^ but this lower dose was not efficacious in two clinical trials.^2,5,7,8^

The ongoing Accelerating Coronavirus Disease 2019 Therapeutic Interventions and Vaccines (ACTIV-6) platform randomized clinical trial evaluates repurposed medications in the outpatient setting.^7^ A previous arm of ACTIV-6 randomized 1331 adults with mild to moderate COVID-19 to receive fluvoxamine 50 mg twice daily or placebo for 10 days.^7^ The primary outcome of time to sustained recovery was not different between the fluvoxamine and placebo groups and no differences were observed in need for higher-level medical care or death. The lack of efficacy with fluvoxamine 50 mg twice daily may be due to an inadequate dose. With conflicting results across several large randomized controlled trials, there is a need to confirm the potential therapeutic benefits of fluvoxamine at the higher 100 mg twice daily dose.

For this study, the ACTIV-6 platform sought to evaluate the effect of higher-dose fluvoxamine (50 mg twice daily for 1 day, followed by 100 mg twice daily for 12 days) on time to sustained recovery from mild to moderate COVID-19 or progression to severe disease in non-hospitalized adults.

## METHODS

### Trial Design and Oversight

ACTIV-6 is a double-blind randomized placebo-controlled platform trial to study repurposed medications for the treatment of outpatients with mild to moderate COVID-19 in the United States.^9^ ACTIV-6 utilizes a decentralized approach for integration into diverse healthcare and community-based settings, including COVID-19 clinical testing and treatment programs. The complete protocol and statistical analysis plan are provided in **Supplement 1**.

The trial protocol was approved by a central institutional review board with review at each site. Informed consent was obtained from each participant either via written or electronic consent. An independent data and safety monitoring committee oversaw participant safety, efficacy, and trial conduct.

### Participants

Since the study platform opened on June 11, 2021, over 7500 participants have been randomized across 6 study arms. The fluvoxamine 100 mg twice daily study arm enrolled participants from 103 sites between August 25, 2022 and January 20, 2023. Participants were identified by individual sites or by self-referral via the central study telephone hotline. The study was closed in anticipation of achieving the prespecified sample size accrual target and due to concerns for limited availability of study product, could not over-enroll.

Study eligibility criteria at the time of screening were confirmed at the site level and included age ≥30 years, SARS-CoV-2 infection confirmed with a positive polymerase chain reaction or antigen test (including home-based testing) within the past 10 days, and actively experiencing ≥2 COVID-19 symptoms for ≤7 days from the time of consent (full eligibility criteria in **Supplement 1**). Symptoms included fatigue, dyspnea, fever, cough, nausea, vomiting, diarrhea, body aches, chills, headache, sore throat, nasal symptoms, and new loss of sense of taste or smell. Individuals meeting the following criteria were excluded from participation: current or recent hospitalization for COVID-19, ongoing or planned participation in other interventional trials for COVID-19, current or recent use (within 14 days) of fluvoxamine or other selective serotonin (or norepinephrine) reuptake inhibitors or monoamine oxidase inhibitors, bipolar disorder, pregnant or nursing, or known allergy or contraindications to fluvoxamine. Prior receipt of COVID-19 vaccinations and current use of approved or emergency use authorization therapeutics for outpatient treatment of COVID-19 were allowed.

### Randomization

Due to the adaptive nature of ACTIV-6, study drugs could be added or removed based on evolving data. Unlike previous active drugs within the platform, the open period of enrollment for fluvoxamine 100 mg did not overlap with the enrollment period of other active drugs. Consequently, the randomization process simplified to a 1:1 matched placebo allocation provided by a random number generator and there was no pooled placebo contribution.

### Interventions

A 13-day supply of either fluvoxamine or matched placebo, provided by the manufacturer (Apotex, Toronto, Canada), was dispensed to the participant via home delivery from a centralized pharmacy. Randomized participants were instructed to self-administer oral fluvoxamine at a dose of 50 mg (one 50 mg tablet) or matching placebo twice daily for 1 day, followed by 100 mg (two 50 mg tablets) or matching placebo twice daily for 12 days, for a total 13-day course.

### Outcome Measures

The primary outcome was time to sustained recovery within 28 days, defined as the time from receipt of drug to the third of 3 consecutive days without COVID-19 symptoms.^7,9^ This measure was selected *a priori* from the 2 possible primary outcomes of the platform. The other possible primary outcome—time to hospitalization or death—transitions to a secondary outcome when not selected as the primary outcome, per the statistical analysis plan. Participants who died within the follow-up period were deemed to have not recovered, regardless of whether they were symptom free for 3 consecutive days. Secondary outcomes included 3 time-to-event endpoints administratively censored at day 28: time to death (number of events permitting), time to hospitalization or death (number of events permitting), or time to first healthcare utilization (a composite of urgent care visits, emergency department visits, hospitalization, or death). Additional secondary outcomes included mean time spent unwell through day 14 and the WHO COVID Clinical Progression Scale on days 7, 14, and 28. Quality of life measures using the PROMIS-29 are being collected through day 180 and are not included in this report.

### Trial Procedures

The ACTIV-6 platform was designed to occur remotely, with all screening and eligibility procedures reported by participants and confirmed at the site level. Positive laboratory results for SARS-CoV-2 were verified by study staff prior to randomization. During screening procedures, participants shared demographic information, medical history, use of concomitant medications, COVID-19 symptoms, and completed quality of life surveys.

A centralized investigational pharmacy packaged and provided active or placebo study products via mail to the address provided by participants. Shipping and delivery information was provided by the courier.

Daily assessments were reported by participants via the study portal during the first 14 days of the study, regardless of symptom status. If participants were not recovered by day 14, the daily assessments continued until recovery or day 28. Planned remote follow-up visits occurred on days 28, 90, and 120. Additional study procedure details are provided in **Supplement 2**.

### Statistical Analysis Plan

Inferences about the primary outcome and exploratory analyses involving secondary outcomes were based primarily on covariate-adjusted regression modeling, supplemented by unadjusted models and graphical and tabular displays. Proportional hazard regression was used for the time-to-event analysis, and cumulative probability ordinal regression models were utilized for ordinal outcomes. Longitudinal ordinal regressions models were used to estimate the differences in mean time spent unwell, a summary which compares the number of days in the first 14 days of follow-up spent unrecovered.

The planned primary endpoint analysis was a Bayesian proportional hazards model. The primary inferential, decision-making quantity was the posterior distribution for the treatment assignment hazard ratio (HR), with a HR>1 indicating a beneficial effect. If the posterior probability of benefit exceeded 0.95 during interim or final analyses, efficacy of the intervention would be met. To preserve type I error <0.05, the prior for the treatment effect parameter on the log relative hazard scale was a normal distribution centered at 0 and scaled to a standard deviation (SD) of 0.1. All other parameter priors were weakly informative, using the software default of 2.5 times the ratio of the SD of the outcome divided by the SD of the predictor variable. The study was designed to have 80% power to detect a HR of 1.2 in the primary endpoint from a total sample size of 1200 participants with planned interim analyses at 300, 600, and 900 participants.

The model for the primary endpoint included the following predictor variables: randomization assignment, age, sex, duration of symptoms prior to study drug receipt, calendar time, vaccination status, geographical location, call center indicator, and baseline symptom severity. The proportional hazard assumption of the primary endpoint was evaluated by generating visual diagnostics such as the log-log plot and plots of time-dependent regression coefficients for each predictor in the model.

Secondary endpoints were analyzed with Bayesian regression models (either proportional hazards or proportional odds). Weakly informative priors were used for all parameters. Secondary endpoints were not used for formal decision-making, and no decision threshold was selected. Analyses resulting from secondary endpoints should be interpreted as exploratory given the potential for type I errors due to multiple comparisons. The same sets of covariates used in the primary endpoint model were used in the analysis of the secondary endpoints, provided that the endpoint accrued sufficient events to be analyzed with covariate adjustment.

All available data were utilized to compare each active study drug vs placebo control, regardless of post-randomization adherence to study protocols. The modified intention-to-treat (mITT) cohort comprised all participants who were randomized, who did not withdraw before delivery of study drug, and for whom the courier confirmed study drug delivery. Day 1 of the study was defined as the day of study drug delivery. Participants who opted to discontinue data collection were censored at the time of last contact, including those participants who did not complete any surveys or phone calls after receipt of study drug. Missing data among covariates for both primary and secondary analyses were addressed with conditional mean imputations because the amount of missing covariate data was small.

A predefined analysis examined potential variations in treatment effects based on participant characteristics. The assessment of treatment effect heterogeneity encompassed age, symptom duration, body mass index (BMI), symptom severity on day 1, calendar time (indicative of circulating SARS-CoV-2 variant), sex, and vaccination status. Continuous variables were analyzed as such, without stratifying into subgroups.

Analyses were performed with R^10^ version 4.3 with the following primary packages: rstanarm,^11,12^ rmsb,^13^ and survival.^14^

## RESULTS

### Study Population

A total of 1208 participants provided consent and were randomized to either fluvoxamine or placebo. A small subset of 33 participants were excluded from the analysis population post randomization because study drug was not delivered within 7 days of randomization. The modified intent to treat (mITT) cohort included the 1175 participants who were randomized, received study drug, and did not withdraw from the study before receiving study drug. In the mITT analysis cohort, 589 participants were randomized to receive fluvoxamine and 586 participants were randomized to placebo (**Figure 1**).

**Figure 1.**
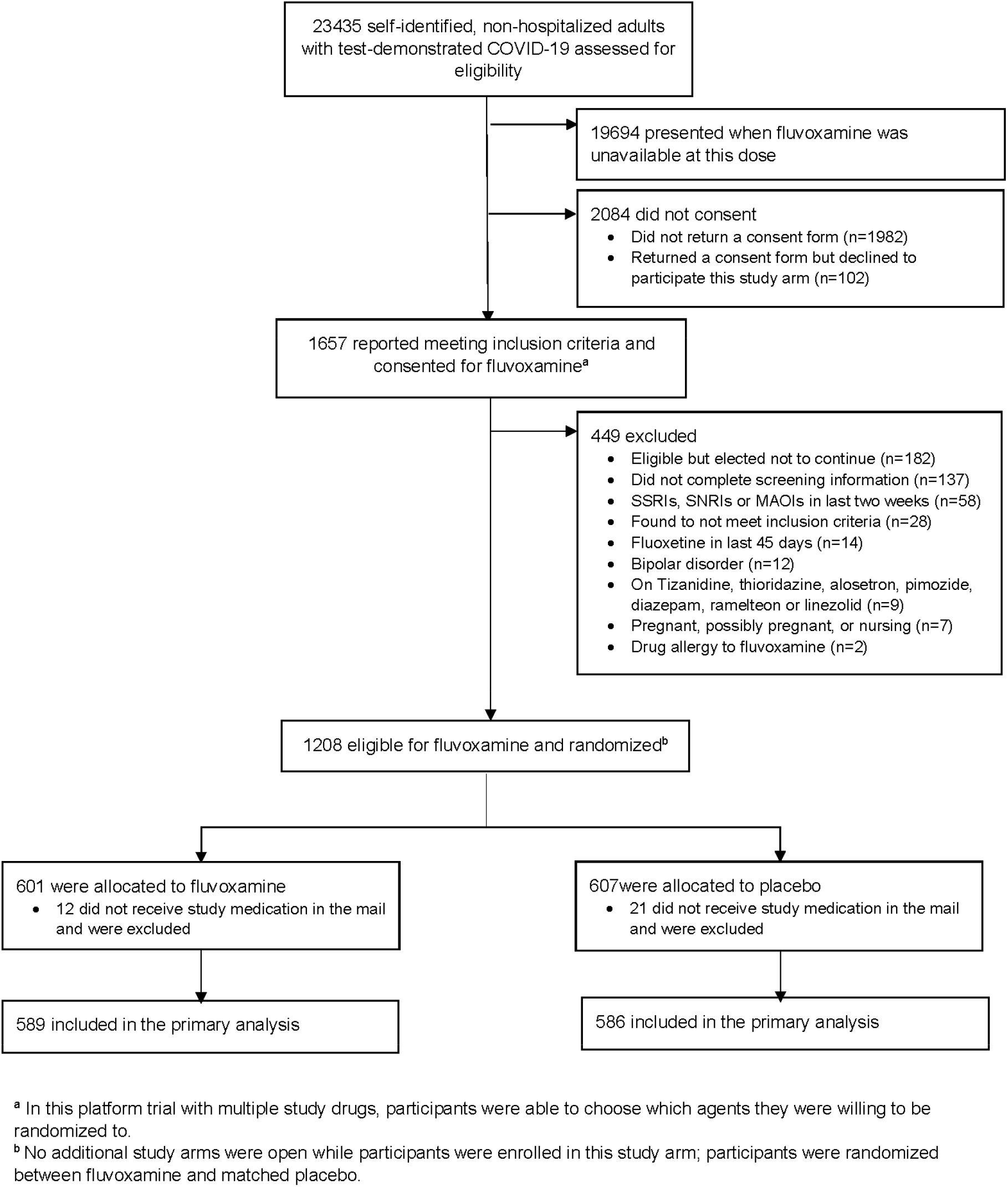
Participant flow in a trial of higher-dose fluvoxamine for mild to moderate COVID-19

The characteristics of the participant population were similar to those observed in other ACTIV-6 cohorts (**Table 1**). The median age was 50 years (IQR 40–60); 66% were female; the most commonly self-reported races were White (73%), Black/African American (9%), Middle Eastern (6%), or Asian (5%), and 45% of participants identified as Hispanic/Latino. The most common comorbidities were obesity (36%) and hypertension (26%). Overall, 77% of participants reported having received at least 2 SARS-CoV-2 vaccine doses. About 13% of participants reported taking a recommended COVID-19 therapy (**Table 1**).

**Table 1.**
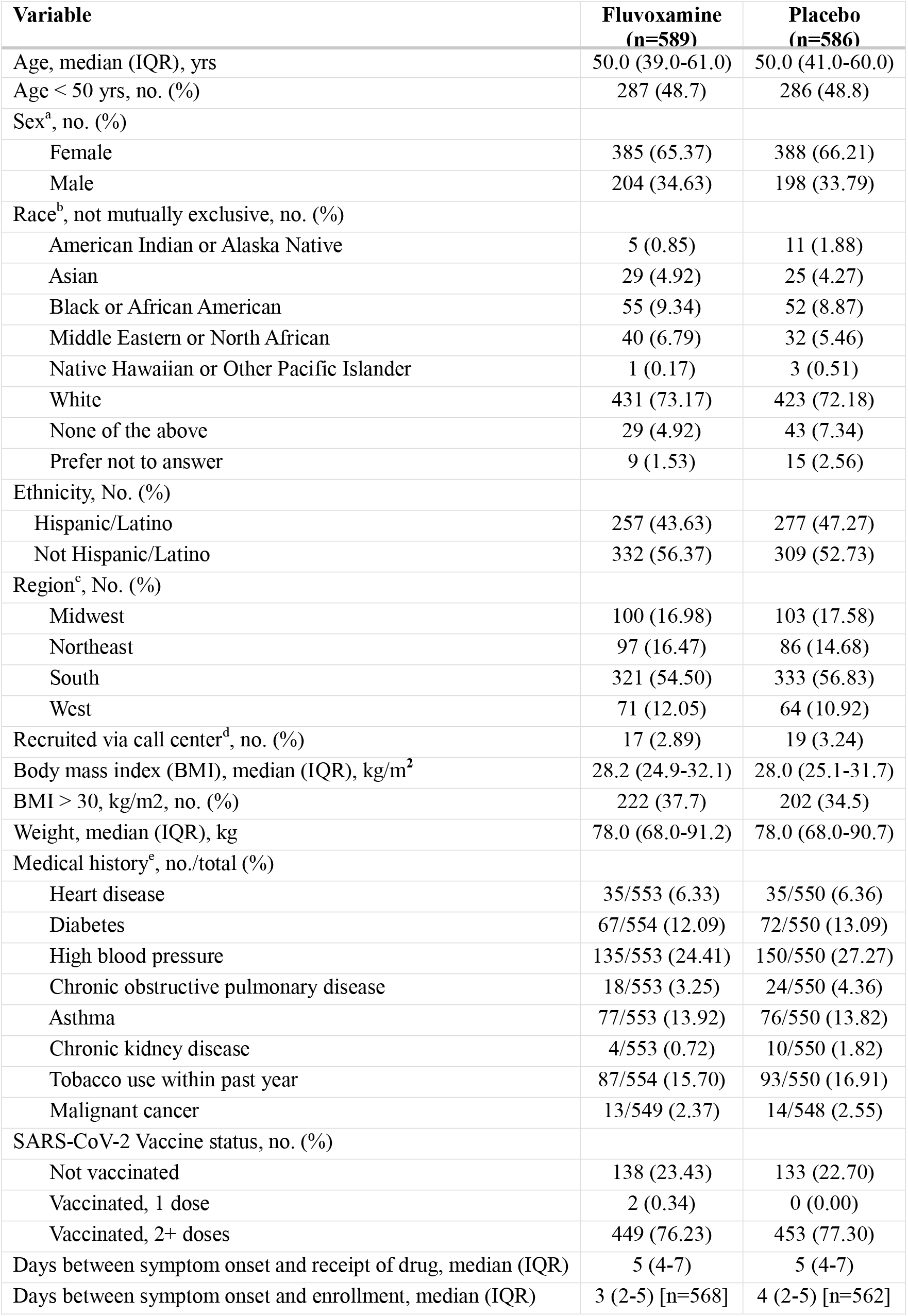
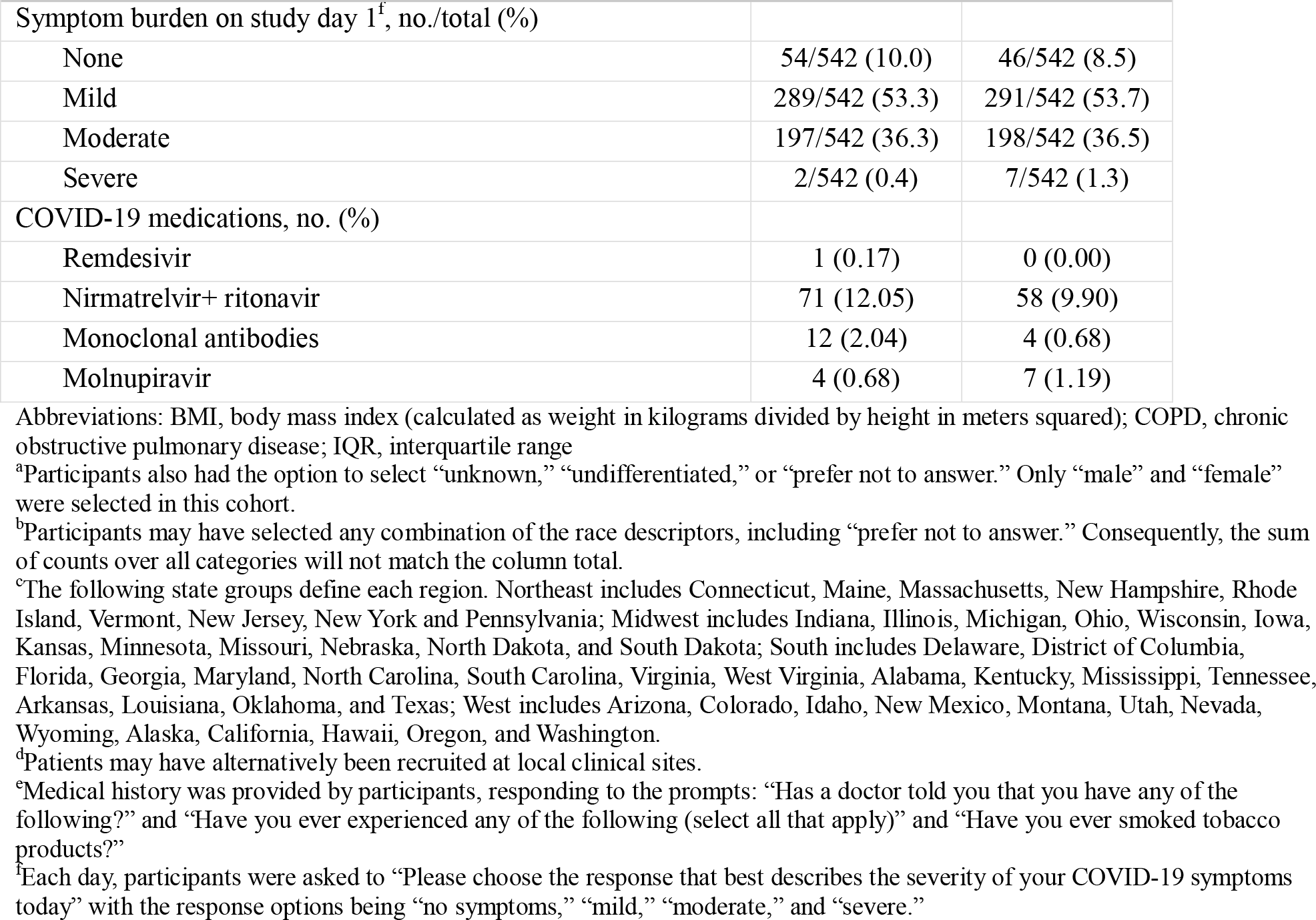
Baseline characteristics.

| Variable | Fluvoxamine<br>(n=589) | Placebo<br>(n=586) |
| --- | --- | --- |
| Age, median (IQR), yrs | 50.0 (39.0-61.0) | 50.0 (41.0-60.0) |
| Age < 50 yrs, no. (%) | 287 (48.7) | 286 (48.8) |
| Sex<sup>a</sup>, no. (%) |  |  |
| Female | 385 (65.37) | 388 (66.21) |
| Male | 204 (34.63) | 198 (33.79) |
| Race<sup>b</sup>, not mutually exclusive, no. (%) |  |  |
| American Indian or Alaska Native | 5 (0.85) | 11 (1.88) |
| Asian | 29 (4.92) | 25 (4.27) |
| Black or African American | 55 (9.34) | 52 (8.87) |
| Middle Eastern or North African | 40 (6.79) | 32 (5.46) |
| Native Hawaiian or Other Pacific Islander | 1 (0.17) | 3 (0.51) |
| White | 431 (73.17) | 423 (72.18) |
| None of the above | 29 (4.92) | 43 (7.34) |
| Prefer not to answer | 9 (1.53) | 15 (2.56) |
| Ethnicity, No. (%) |  |  |
| Hispanic/Latino | 257 (43.63) | 277 (47.27) |
| Not Hispanic/Latino | 332 (56.37) | 309 (52.73) |
| Region<sup>c</sup>, No. (%) |  |  |
| Midwest | 100 (16.98) | 103 (17.58) |
| Northeast | 97 (16.47) | 86 (14.68) |
| South | 321 (54.50) | 333 (56.83) |
| West | 71 (12.05) | 64 (10.92) |
| Recruited via call center<sup>d</sup>, no. (%) | 17 (2.89) | 19 (3.24) |
| Body mass index (BMI), median (IQR), kg/m<sup>2</sup> | 28.2 (24.9-32.1) | 28.0 (25.1-31.7) |
| BMI > 30, kg/m<sup>2</sup>, no. (%) | 222 (37.7) | 202 (34.5) |
| Weight, median (IQR), kg | 78.0 (68.0-91.2) | 78.0 (68.0-90.7) |
| Medical history<sup>e</sup>, no./total (%) |  |  |
| Heart disease | 35/553 (6.33) | 35/550 (6.36) |
| Diabetes | 67/554 (12.09) | 72/550 (13.09) |
| High blood pressure | 135/553 (24.41) | 150/550 (27.27) |
| Chronic obstructive pulmonary disease | 18/553 (3.25) | 24/550 (4.36) |
| Asthma | 77/553 (13.92) | 76/550 (13.82) |
| Chronic kidney disease | 4/553 (0.72) | 10/550 (1.82) |
| Tobacco use within past year | 87/554 (15.70) | 93/550 (16.91) |
| Malignant cancer | 13/549 (2.37) | 14/548 (2.55) |
| SARS-CoV-2 Vaccine status, no. (%) |  |  |
| Not vaccinated | 138 (23.43) | 133 (22.70) |
| Vaccinated, 1 dose | 2 (0.34) | 0 (0.00) |
| Vaccinated, 2+ doses | 449 (76.23) | 453 (77.30) |
| Days between symptom onset and receipt of drug, median (IQR) | 5 (4-7) | 5 (4-7) |
| Days between symptom onset and enrollment, median (IQR) | 3 (2-5) [n=568] | 4 (2-5) [n=562] |
| Symptom burden on study day 1<sup>f</sup>, no./total (%) |  |  |
| None | 54/542 (10.0) | 46/542 (8.5) |
| Mild | 289/542 (53.3) | 291/542 (53.7) |
| Moderate | 197/542 (36.3) | 198/542 (36.5) |
| Severe | 2/542 (0.4) | 7/542 (1.3) |
| COVID-19 medications, no. (%) |  |  |
| Remdesivir | 1 (0.17) | 0 (0.00) |
| Nirmatrelvir+ ritonavir | 71 (12.05) | 58 (9.90) |
| Monoclonal antibodies | 12 (2.04) | 4 (0.68) |
| Molnupiravir | 4 (0.68) | 7 (1.19) |

Abbreviations: BMI, body mass index (calculated as weight in kilograms divided by height in meters squared); COPD, chronic obstructive pulmonary disease; IQR, interquartile range
<sup>a</sup>Participants also had the option to select “unknown,” “undifferentiated,” or “prefer not to answer.” Only “male” and “female” were selected in this cohort.
<sup>b</sup>Participants may have selected any combination of the race descriptors, including “prefer not to answer.” Consequently, the sum of counts over all categories will not match the column total.
<sup>c</sup>The following state groups define each region. Northeast includes Connecticut, Maine, Massachusetts, New Hampshire, Rhode Island, Vermont, New Jersey, New York and Pennsylvania; Midwest includes Indiana, Illinois, Michigan, Ohio, Wisconsin, Iowa, Kansas, Minnesota, Missouri, Nebraska, North Dakota, and South Dakota; South includes Delaware, District of Columbia, Florida, Georgia, Maryland, North Carolina, South Carolina, Virginia, West Virginia, Alabama, Kentucky, Mississippi, Tennessee, Arkansas, Louisiana, Oklahoma, and Texas; West includes Arizona, Colorado, Idaho, New Mexico, Montana, Utah, Nevada, Wyoming, Alaska, California, Hawaii, Oregon, and Washington.
<sup>d</sup>Patients may have alternatively been recruited at local clinical sites.
<sup>e</sup>Medical history was provided by participants, responding to the prompts: “Has a doctor told you that you have any of the following?” and “Have you ever experienced any of the following (select all that apply)” and “Have you ever smoked tobacco products?”
<sup>f</sup>Each day, participants were asked to “Please choose the response that best describes the severity of your COVID-19 symptoms today” with the response options being “no symptoms,” “mild,” “moderate,” and “severe.”

While only participants with symptoms were enrolled in the study, some participants reported no symptoms on the day of study drug delivery, which is defined as study day 1. At the time of study drug delivery, 9% of participants reported no symptoms, while the majority reported mild (54%) or moderate (36%) symptoms. The symptom burden for each of the 13 COVID19-related symptoms at baseline is reported in **eTable 1**. Participants were enrolled within a median of 3 days of patient-reported symptom onset (IQR 2–5 days) and study drug was delivered within a median of 5 days of symptom onset (IQR 4– 7 days). The complete distribution of time between onset of COVID-19 symptoms and study drug delivery is reported in **eFigure 1**.

### Primary Outcome

Differences in time to sustained recovery were not observed in either unadjusted Kaplan-Meier curves (**Figure 2**) or covariate-adjusted regression models (**Table 2**). The median time to sustained recovery was 10 days (95% CI 10-11) in both the fluvoxamine and placebo groups. The posterior probability for benefit was 0.4, with an HR of 0.99 (95% CrI 0.89–1.09) (**Figure 3**). Sensitivity analyses yielded similar estimates of the treatment effect (**eFigure 2**).

**Table 2.**
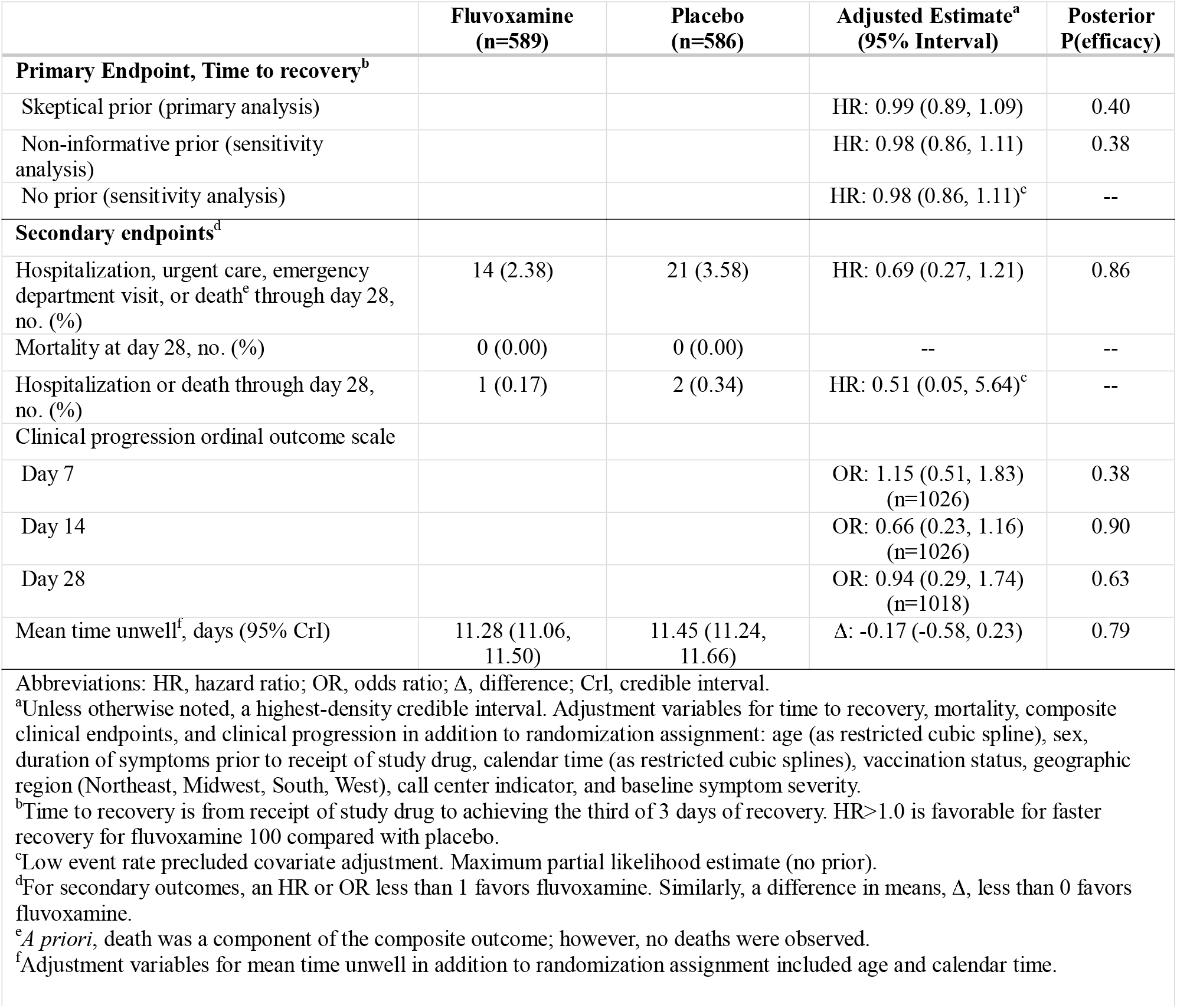
Primary and Secondary Outcomes.

**Figure 2.**
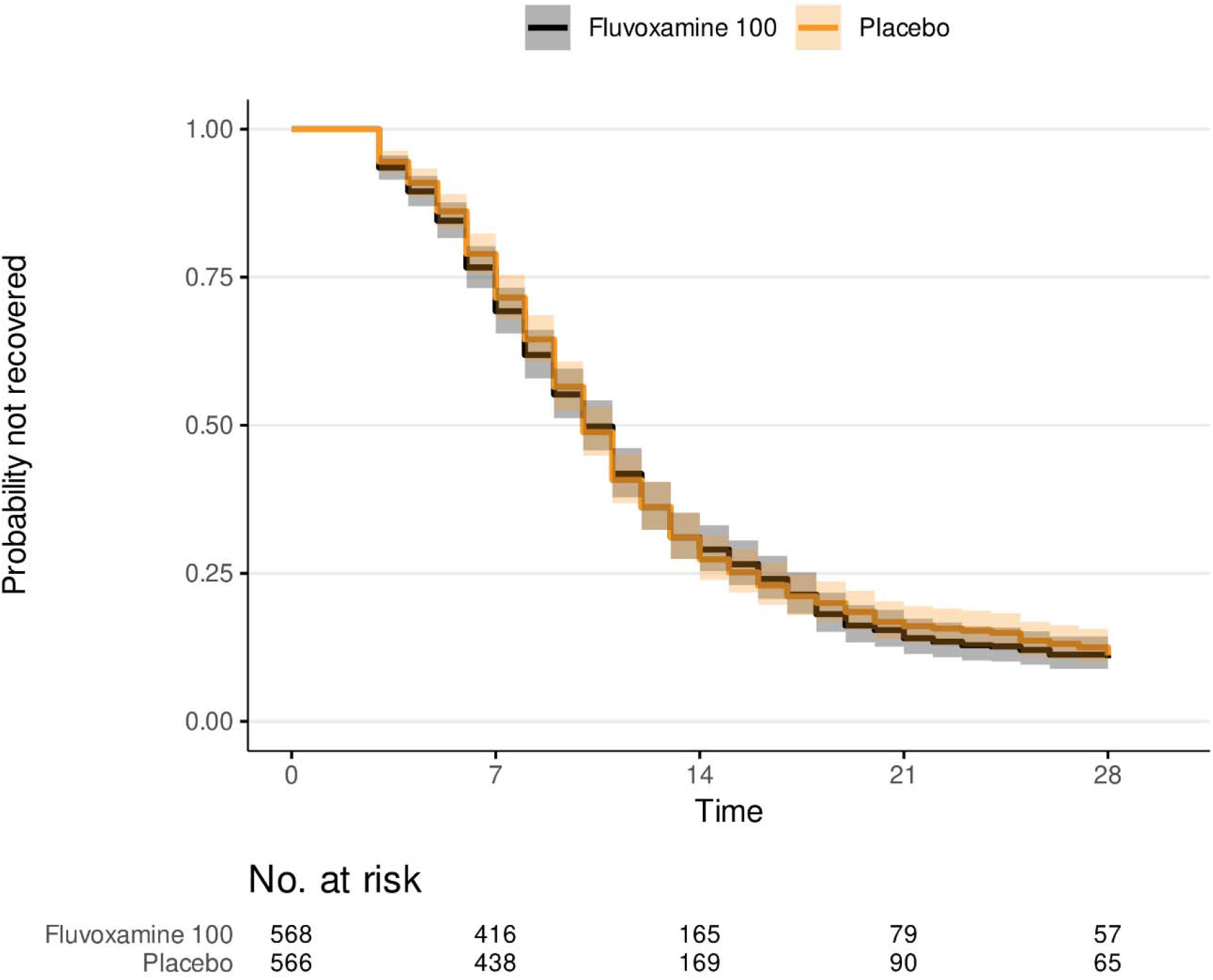
Primary outcome of time to sustained recovery Recovery was defined as the third of 3 consecutive days without symptoms. Fourty-five participants were censored for complete nonresponse, 51 were censored after partial response, and all others were followed up until recovery, death, of the end of short-term 28-day follow-up. Median time to recovery was 10 (95% CI 10 to 11) days in the fluvoxamine and placebo groups. Shaded regions denote the pointwise 95% CIs.

**Figure 3.**
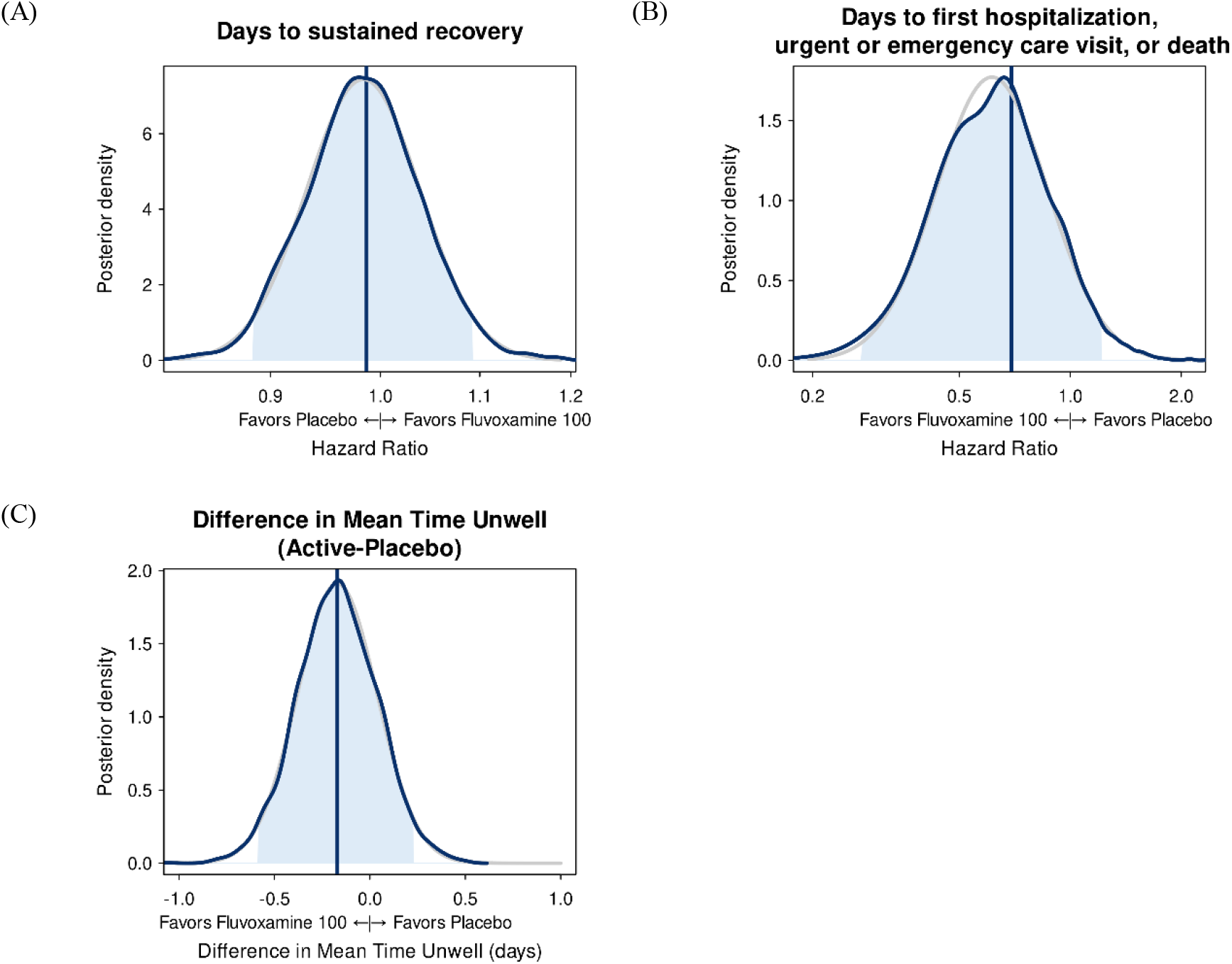
Time to sustained recovery; hospitalization, urgent or emergency care visits, or death; and mean time unwell The vertical lines represent the estimated mean of the posterior distribution. Posterior density is the relative likelihood of posterior probability distribution. Outcomes with higher posterior density are more likely than outcomes with lower posterior density. Blue density lines are kernel density estimates constructed from posterior draws. Grey density lines are parametric estimates, also estimated from posterior draws. The posterior density plots of all the covariates in the primary outcome model are shown in the supplement (**eFigure 9**).

### Secondary Outcomes

No deaths were observed in either group; 1 participant in the fluvoxamine group and 2 in the placebo group were hospitalized (**Table 2, eFigure 3**). There were 14 (2.4%) participants in the fluvoxamine group and 21 (3.6%) in the placebo group who reported hospital admission or emergency department or urgent care visits (**Table 2, eFigure 4**). Analyzed as a time to first event outcome, the HR for the composite healthcare outcome was 0.69 (95% CrI 0.27–1.21) with a posterior probability of efficacy of 0.86 (**Figure 3**).

With clinical events like hospitalization and death being rare among participants, the COVID clinical progression scale (**Supplement 2**) simplified into a self-reported evaluation of home activity levels (limited vs not) collected on study days 7, 14, and 28 (**eFigure 5**). By day 7, more than 95% of responding participants reported no limitations in activity, thus this endpoint did not meet prespecified thresholds for beneficial treatment effect. Likewise, the difference in mean time unwell was similar between the fluvoxamine and placebo groups (11.3 days [CI 11.1–11.5] vs 11.5 days [CI 11.2–11.7]; 95% CrI -0.58 to 0.23; P[efficacy]=0.79). (**Figure 3**)

### Adverse Events and Tolerability

Seven serious adverse events were reported in six participants (**eTable 2**), all among participants who reported taking study medication at least once. Three events in two participants were reported in the fluvoxamine group, including aggravated asthma, community-acquired pneumonia, and Guillain-Barre syndrome. Four participants in the placebo group reported one serious adverse event each, including ruptured appendix, diabetic foot ulcer, partial bowel obstruction, and perforated intestinal diverticulitis. The incidence of serious events was low without evidence of higher events in the intervention group.

Among participants who reported adherence data, 6.4% (36/561) in the fluvoxamine group compared with 2.1% (13/563) in the placebo group reported at least once “I am not planning to take my medicine because I feel worse,” consistent with a prior hypothesis that fluvoxamine may not be universally tolerated, especially at higher doses.

### Heterogeneity of Treatment Effect Analyses

When stratified by baseline symptom severity and the timing of treatment relative to the onset of symptoms, no meaningful separation between fluvoxamine and placebo participants was observed in the distribution of the primary outcome (**eFigure 6**). Likewise, exploratory analyses to understand how the treatment effect for the primary outcome may vary with a priori defined patient characteristics was completed. The analysis suggests the possibility that participants who received fluvoxamine sooner after symptom onset faired poorer than placebo (**eFigure 7**); whereas, participants who received fluvoxamine about 7 days after symptom onset may have done better than placebo (interaction p-value: 0.05). In additional exploratory analyses without covariate adjustment, the differences in recovery between those that received drug within 3 days of symptom onset and those that received drug on days 6 to 8 were plotted (**eFigure 8**).

## DISCUSSION

Among outpatient adults with mild to moderate COVID-19, treatment with fluvoxamine 100 mg twice daily for 13 days, compared with placebo, did not improve time to sustained recovery in this large trial of 1175 participants. The present study is one in a series of trials investigating fluvoxamine as a potential treatment for COVID-19 in an outpatient setting. STOP COVID^5^ (n=152), TOGETHER^6^ (n=1497), and COVID OUT^2^ (n=661) trials were designed around clinical events such as death, hospitalization, hypoxemia, and emergency department visits. The TOGETHER trial was stopped early for superiority of fluvoxamine 100 mg twice daily, compared with placebo, with a 32% reduction in the primary composite endpoint of hospitalization or extended care in an emergency setting.^5^ A follow up TOGETHER trial (n=1476) testing fluvoxamine 100mg twice daily with inhaled budesonide in a majority vaccinated population demonstrated a 50% reduction in the same composite endpoint.^8^ In contrast, the COVID-OUT and ACTIV-6 trials studying fluvoxamine 50 mg twice daily did not identify a benefit for fluvoxamine.^2^ ACTIV-6 is the only trial with a primary outcome of patient-reported sustained recovery, and in this trial of fluvoxamine 100 mg twice daily for 13 days, there was no evidence of benefit. A secondary composite outcome of death or healthcare utilization (including urgent care or emergency department visits or hospitalizations) suggested one-third fewer events in the fluvoxamine group compared with placebo. Although this difference was not conclusive at the defined decision-making thresholds, the clinical value of reducing disease progression and the need for healthcare utilization should not be underestimated from the patient’s perspective.

The evolution of the pandemic, with changes in the severity of COVID-19 over time and increasing natural and/or vaccine-induced immunity, suggests that the circumstances of the present study are meaningfully different than those of earlier trials. COVID-19 severity has abated over time with lower rates of hospitalization and death as partial immunity has increased. If clinical event rates for death or hospitalizations continue to decline, future trials may need to focus on composite outcomes such as healthcare utilization or enroll much larger number of participants to conclusively understand the impact of any therapeutic agent on outcomes. Testing time to recovery in clinical trials remains appealing as a symptom reduction is clinically relevant and patient-centric; however, several medications including nirmatrelvir, metformin, and molnupiravir do not reduce symptom duration while showing clinical benefit.^2,15^

Exploratory findings from the heterogeneity of treatment effect analysis prompt the hypothesis that participants receiving fluvoxamine later in the course of their COVID-19 infection may have had more rapid symptom resolution compared to placebo in contrast to the difference between those who received drug earlier in the course of disease. A plausible hypothesis is that the immune modulating activity of fluvoxamine may not be beneficial until the latter stage of disease when the host experiences dysregulated immune responses.^16^

A strength of this study is the improved ethnic diversity, with 45% of participants identifying as Hispanic/Latino ethnicity. The pandemic exposed significant health disparities, with a large burden of more severe disease outcomes being reported from underrepresented and marginalized populations including Black/African American and Hispanic/Latino communities.^17–19^ This increased diversity in ACTIV-6 came from a concerted national effort to improve recruitment strategies, prioritize Spanish language document translation, and onboard sites from diverse communities.

### Limitations

The trial has several limitations. Due to the evolving nature of the pandemic and the population enrolled, few clinical events occurred, limiting the ability of the trial to study the treatment effect on clinical outcomes either as a primary or secondary outcomes. The remote nature of the trial, while in part a strength, is also a limitation. The decentralized approach in theory expands access by allowing participation regardless of geographic location. The primary limitation of the remote design is that study drug must be sent by courier, which resulted in additional days between symptom onset and the start of treatment, which could be particularly relevant for a proposed antiviral mechanism of action.

## Conclusions

Among outpatient adults with mild to moderate COVID-19, treatment with fluvoxamine 100 mg twice daily, compared with placebo, did not improve time to sustained recovery. While one-third fewer healthcare utilization events occurred in the fluvoxamine intervention group, the difference was not conclusive.

## Data Availability

Prior to deposition of the data in a public repository which will occur when the platform trial has concluded, persons may request the data by submitting a proposal. If the data can be used for the proposed purpose and it is consistent with the informed consent, then the data will be released under a Data Use Agreement.

## Funding/Support

ACTIV-6 is funded by the National Center for Advancing Translational Sciences (NCATS) (3U24TR001608-06S1). Additional support for this study was provided by the Office of the Assistant Secretary for Preparedness and Response, Biomedical Advanced Research and Development Authority (Contract No.75A50122C00037). The Vanderbilt University Medical Center Clinical and Translational Science Award from NCATS (UL1TR002243) supported the REDCap infrastructure.

## Group Information

The members of the Accelerating COVID-19 Therapeutic Interventions and Vaccines (ACTIV)-6 Study Group and Investigators appear in the Supplement.

## Notes

### Competing Interest Statement

Authorship is attributed to the ACTIV-6 Study Group. Funding information for the ACTIV-6 study is included in the manuscript.

### Clinical Trial

ClinicalTrials.gov (NCT04885530).

### Author Declarations

WCG IRB gave ethical approval for this work.

